# Transjugular Intrahepatic Portosystemic Shunt and Locoregional Therapies in Patients Undergoing Orthotopic Liver Transplantation: A Protocol for a Retrospective, Linked United Network for Organ Sharing Cohort

**DOI:** 10.1101/2021.09.12.21263391

**Authors:** Menelaos Konstantinidis, John T. Moon, Peiman Habibollahi, Hyun S. Kim, Minzhi Xing, Nariman Nezami

**Author notes:** **Corresponding authors:** Nariman Nezami, MD, 22 S. Greene Street, Baltimore MD 21201, U.S.A. **First authors:** MK and JTM are equal contributors to this article.

## Abstract

**Introduction:** Orthotopic Liver Transplantation (OLT) is the potential curative treatment option for patients with end-stage liver disease (ESLD) or hepatocellular carcinoma (HCC) within organ procurement and transplantation network (OPTN) criteria. However, these groups of patients may require bridging interventions, including Transjugular Intrahepatic Portosystemic Shunt (TIPS) or Locoregional Therapies (LRTs), given the nationwide organ shortage and increasing waitlist time. The perioperative and long-term post-OLT survival and clinical outcomes require further investigation to evaluate the clinical utility and therapeutic advantages of these bridging interventions, if any. We propose a large retrospective database analysis that will evaluate both perioperative and long-term effects of these OLT-related interventions.

**Methods and analysis:** Three datasets from the United Network for Organ Sharing (UNOS) database will be included and linked to estimate the causal effect of 1) Transjugular Intrahepatic Portosystemic Shunts and 2) Locoregional therapies in patients undergoing OLT, the latter among patients with HCC. Only therapy naïve adult patients, without multivisceral transplants, and without living donor transplants will be included. The primary outcome will be overall survival. Secondary outcomes will include perioperative clinical outcomes, post-operative survival, and postoperative clinical outcomes. The inverse probability of treatment weighted models with Cox regression will be utilized to analyze survival outcomes, logistic regression for categorical outcomes, and ordinary least squares regression for continuous outcomes. A sensitivity analysis will be conducted to assess the appropriateness of a complete-case analysis for the primary outcome and ensure the robustness of the findings.

**Ethics and Dissemination:** This study protocol was reviewed by the Emory University School of Medicine Institutional Review Board (IRB), and ethical approval was waived due to the retrospective analysis of the originally anonymized database. The results will be disseminated in peer-reviewed journals and presented at relevant conferences. It was not appropriate or possible to involve patients or the public in the design, or conduct, or reporting, or dissemination plans of our research.

**STRENGTHS AND LIMITATIONS OF THIS STUDY:** *Strengths:* The proposed study: - Will be the first study evaluating the causal effect of TIPS in OLT candidates and of locoregional therapies in OLT candidates with HCC
- Will be the first study to link UNOS datasets to investigate the estimands, thereby providing insight into the clinical impact of TIPS and LRTs at various stages in the clinical pathway.

*Limitations:* The proposed study: - Will be a retrospective study and thus subject to poor or inadequate reporting in the registry, though propensity score matching will be done
- May be subject to unmeasured confounding and sensitive to model misspecification
- May lack the necessary sample size and subsequently be underpowered to estimate the target estimands

## INTRODUCTION

End-stage liver disease (ESLD) is often the product of chronic liver disease which regardless of its etiology, can lead to cirrhosis and portal hypertension (PHT) [1]. Portal hypertension results in high risk of recurrent esophageal varices, gastric variceal bleeding, and paracentesis-refractory ascites [2, 3]. If any of these complications of PHT happens, Transjugular Intrahepatic Portosystemic Shunt (TIPS) could play an important and effective role in risk mitigation by relieving portal hypertension and its related downstream effects [4].

Cirrhotic patients are also at a higher risk for developing hepatocellular carcinoma (HCC), one of the most common oncologic diseases with approximately 500,000 diagnoses annually worldwide [5, 6]. With or without HCC, Orthotopic Liver Transplant (OLT) is the definitive therapy for ESLD [7]. However, given the rising demand and persistent liver donor shortage despite implementing different transplantation policies, many of the existing treatments are aimed at successfully bridging ESLD patients to transplantation or curbing donor waiting-list mortality [8, 9]. Those with HCC may be candidates to receive locoregional therapy (LRT), which encompasses a variety of therapeutic interventions, including percutaneous ablations such as radiofrequency ablation (RFA), cryoablation (Cryo), microwave ablation (MVA), or transarterial approaches such as bland embolization, chemoembolization, and radioembolization [10]. The survival effect and clinical impact of TIPS and LRT in ESLD patients pre- and post-OLT has not been well-studied and is the topic of our investigation.

### Importance

The long-term survival outcomes and clinical impact of TIPS in the post-OLT setting have not been well studied for patients with and without HCC. Similarly, the effect of LRT or LRT modality in those with HCC has not been characterized with regard to TIPS placement. Understanding these effects and outcomes will help clinicians to screen and enroll ESLD patients to TIPS as well as to one modality of LRT versus another in patients with HCC.

This work aims to evaluate the effect of TIPS and LRT (separately) on the standard of care for orthotopic liver transplant patients with and without HCC. In particular, the following objectives are of interest:

#### Objective 1

To assess the effect of TIPS on the survival and clinical outcomes of OLT patients with and without HCC

#### Objective 2

To assess the effect of LRTs on the survival and clinical outcomes of OLT patients with HCC

## METHODS

### Study Design and Population

This is a population-based record linkage study of all patients registered in the United Network for Organ Sharing (UNOS) untill March 15, 2021.

Due to the multifactorial nature of the objectives for this study, multiple sets of unique patient populations were defined, which each will be analyzed individually (see below). In detail, our populations are defined as follows:

- Non-HCC patients identified in the UNOS database eligible for TIPS
- HCC patients identified in the UNOS database eligible for TIPS
- HCC patients who had an exception as defined by the policies of the UNOS [11]

Patients who meet the following criteria will be included: adult patients (age>18 years old) and registered in the UNOS waitlist for an organ. Following patients will be excluded: pediatric patients (< 18 years old), patients with OPTN stage 1 (no evidence of HCC on good quality, appropriate surveillance exam), patients who received multivisceral transplants, patients who received living donor transplants, and patients who have already had TIPS or LRTs (as relevant to the corresponding questions) (See Figure 1 for the proposed flow diagram of patient populations and exclusion criteria).

**Figure 1.**
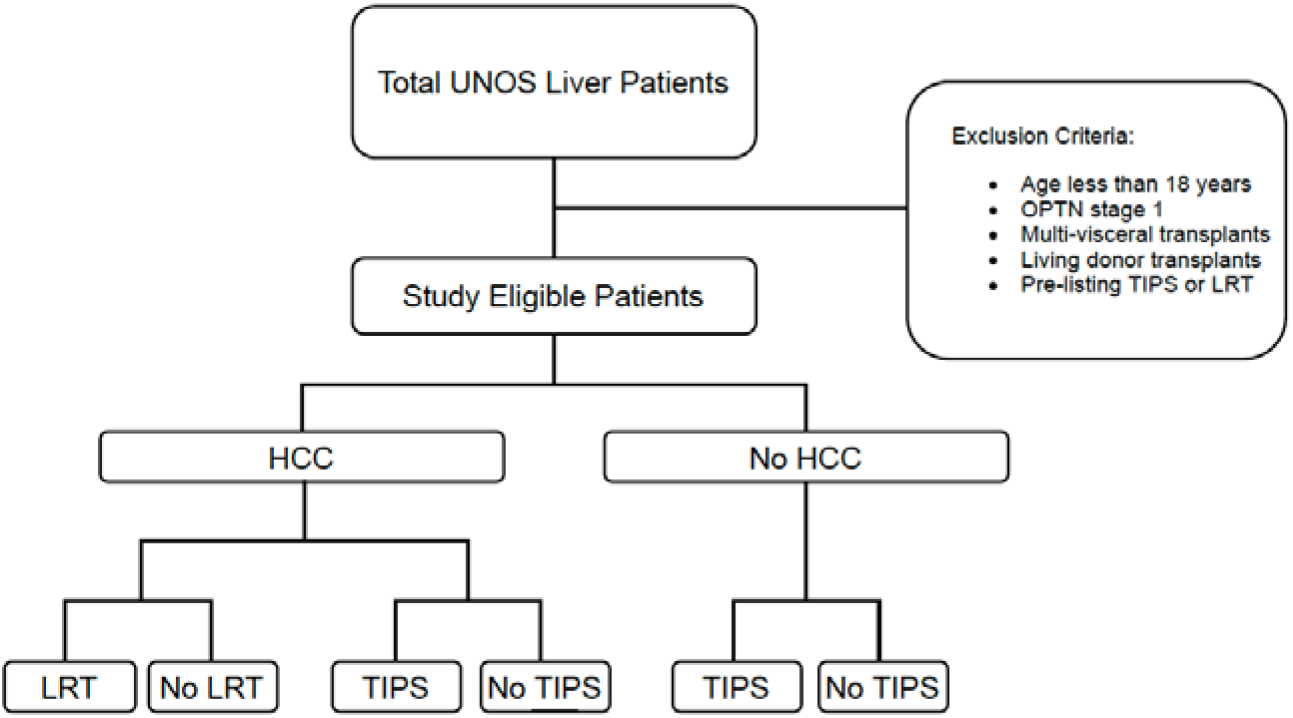
Proposed flow diagram with pre-determined eligibility criteria.

### Overview of the data

The UNOS dataset is a population-based registry of organ transplant patients in the United States. The primary dataset in the present work is the UNOS Liver (UNOSL) dataset, consisting of 321,267 patients from inception to March 15, 2021. On a per-outcome basis, the UNOSL will be linked with the UNOS Liver Follow-up Data (UNOSLF) dataset and the UNOS Liver Exception (UNOSLE) dataset, each consisting of a subset of the UNOSL dataset.

The UNOSL data will provide information on the overall survival of patients, TIPS status, HCC status, and covariates (either at baseline or time-varying). The UNOSLF dataset will provide information on the survival of patients at each follow-up, clinical outcomes, and possibly additional covariates. Lastly, the UNOSLE dataset will provide information on the LRTs undergone by HCC patients (including the modality and number of sessions) and additional covariates such as AFP and the Milan score criteria [12].

### Study Variables

#### Outcomes

For each of the three target populations outlined above, a single primary outcome and several secondary outcomes will be investigated (Table 1). Specifically, the primary outcome of interest will be overall patient survival. The secondary outcomes can be conceptualized into three categories:

**Table 1.**
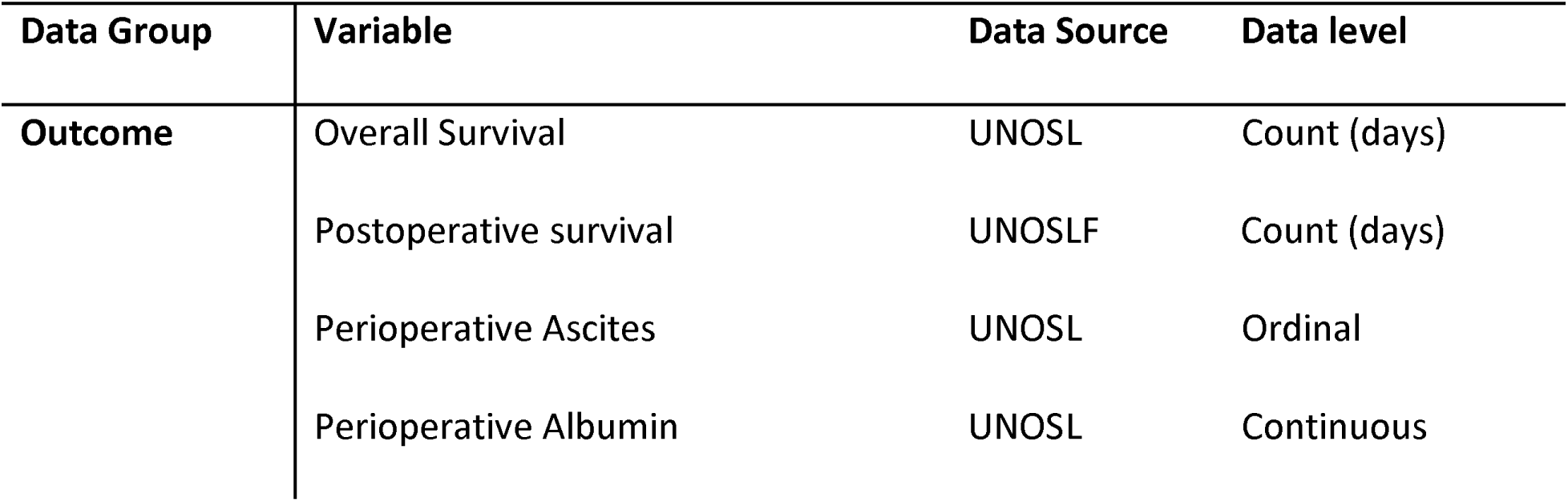
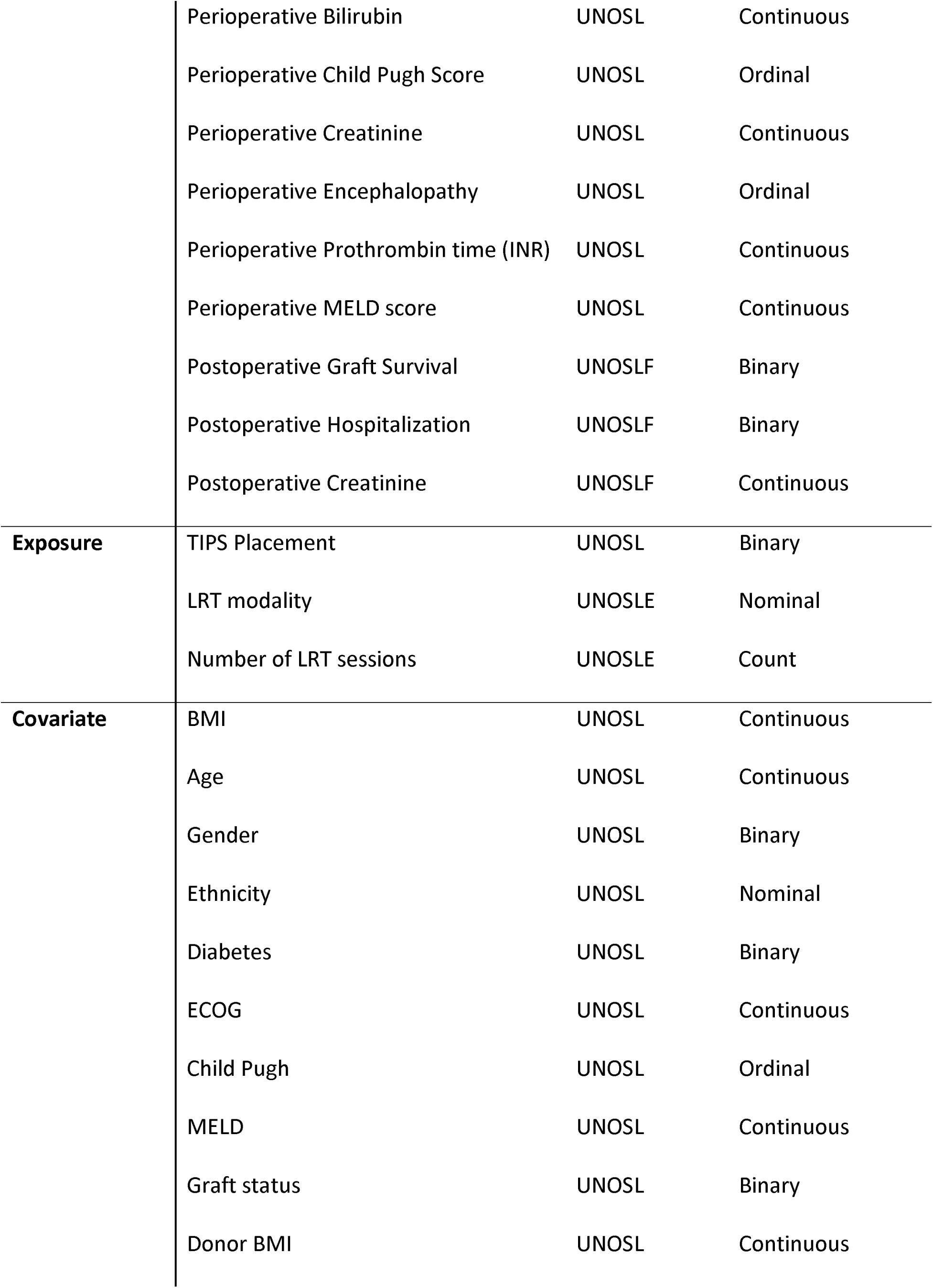
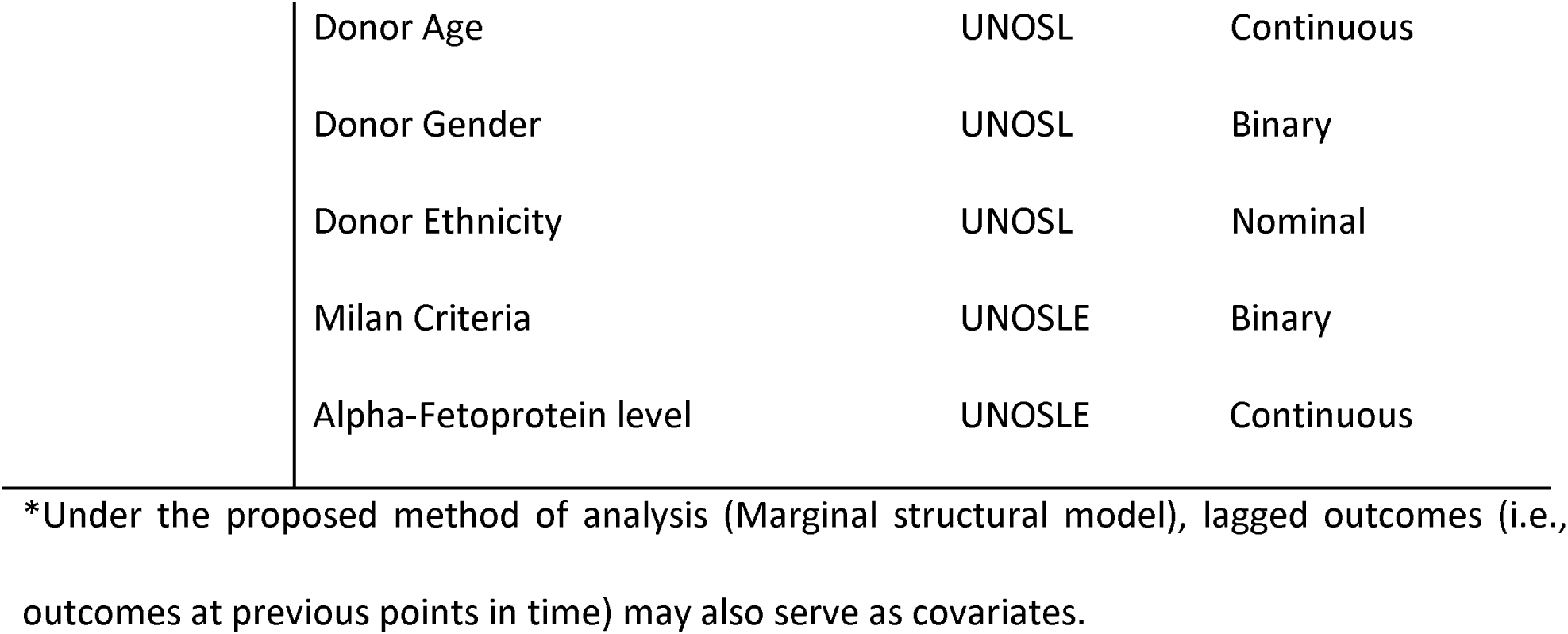
Description of data group, corresponding variables, data source, and variable type to be used in this study

- Perioperative clinical status (Ascites, Encephalopathy, Albumin, Bilirubin, Prothrombin time (expressed using the international normalized ratio [INR], Child Pugh Score, Creatinine (Cr), Eastern Cooperative Oncology Group [ECOG] performance score, and Mayo End-State Liver Disease (MELD) score)
- Post-operative clinical outcomes
- Post-operative survival at 1 month, 3 months, 9 months, 1 year, 2 years, 5 years, and 10-year follow-up

#### Exposures

Throughout, three exposures will be considered, namely, 1) the insertion of TIPS, 2) the type of LRT applied, and 3) the number of LRT sessions. As mentioned in the eligibility criteria above, patients who received multiple types of LRTs will be included in the analyses only if covariates are measured at corresponding time points to allow for time-varying confounding. Otherwise, this will qualify as an additional exclusion criterion, and only patients who underwent a single type of LRT will be included.

#### Follow-ups

For all secondary survival outcomes, linked patients will be included in our study until expiration, leaving the study, or being censored due to the data collection cut-off date (March 15, 2021). In the case of treated HCC patients who underwent OLT, diagnosis of another cancer post-transplant will be an additional exclusion criterion (if the necessary information for making the evaluation is available) when analyzing post-transplant outcomes to avoid biases from competing risks (i.e., death due to underlying condition and death due to cancer). Additionally, it is expected that cause-of-death will be inadequately reported, thus precluding, and possibly biasing, any subsequent competing risk analysis.

#### Covariates

The key covariates will be extracted at baseline (registration) and over time for the proposed analyses. If this is not be possible, the covariate values at baseline will only be extracted. In particular, The aim will be to obtain demographic and patient characteristics including patient Body Mass Index (BMI), age, gender, ethnicity, diagnosis of diabetes, functional status (defined by the Eastern Cooperative Oncology Group [ECOG] performance score) [13], Child Pugh score (i.e., Child-Pugh score)[14], MELD model score [15], graft status, donor BMI, age, gender, and ethnicity, and where applicable (i.e., in the LRT analyses) the Milan Criteria for Liver Transplantation, and Alpha-Fetoprotein level.

### Data linking process

For primary and perioperative outcomes, the UNOSL and UNOSLE datasets will be linked. Patients will be uniquely identified from the UNOSL dataset, and where applicable, the subset of them who are identified in the UNOSLE database will be included for final analysis. In the case of non-HCC patients, only the UNOSL dataset will be used. For post-transplant outcomes, the UNOSL patients will be linked to the UNOSLF patients and, where applicable, to the UNOSLE patients. The set of linkages are summarized in Table 2.

**Table 2.** Summary of data linkage for each outcome and population

| Outcome | Population | UNOSL | UNOSLE | UNOSLF |
| --- | --- | --- | --- | --- |
| Overall Survival | Non-HCC TIPS | X |  |  |
|  | HCC TIPS | X | X |  |
|  | HCC LRT | X | X |  |
| Perioperative Outcomes | Non-HCC TIPS | X |  |  |
|  | HCC TIPS | X | X |  |
|  | HCC LRT | X | X |  |
| Postoperative Outcomes | Non-HCC TIPS | X |  | X |
|  | HCC TIPS | X | X | X |
|  | HCC LRT | X | X | X |

\* 'X' Indicates if a given dataset from UNOSL, UNOSLE, and UNOSLF was linked for a given outcome and population

### Missing data

At present, the extent of the data will be missing is unclear. Although it is not possible to prove if a variable is missing completely at random (MCAR), clinical judgement and past works will be used to evaluate the most likely missing data mechanism (i.e., MCAR as opposed to Missing-at-random or Not-missing-at-random). Should values be judged to be missing completely at random (MCAR), complete case analysis will be proceeded; an analysis in which participants with missing values are excluded from analyses since, under the assumption of MCAR, such analysis will yield unbiased estimates of the treatment effects [16]. For completeness, where MAR or MCAR is plausible, multiple imputations and conduct a sensitivity analysis will be conducted to evaluate the suitability of the complete-case analysis. If a complete-case analysis is found to be appropriate [16-18], this will be reported.

### Statistical Analysis

The descriptive statistics using mean, standard deviation, and quartiles for continuous outcomes, and proportions for categorical variables will be reported. Descriptive statistics will be presented for each treatment stratum of our three target populations (defined above). As per existing recommendations [19], these results will be presented descriptively without corresponding confidence intervals or p-values.

For the primary outcomes, the effect of the exposures will be estimated by weighted multivariable cox regression. If covariates are measured sufficiently measured (e.g., at regular intervals), then the analysis will be conducted through a marginal structural model [20-23] a well-established causal framework for observational studies. If this is not possible, the models through the inverse probability of treatment weights (IPTW) will be implemented [24, 25], defined with respect to the covariates measured at baseline. While not accounting for time-varying confounding, the latter option, subject to the satisfaction of underlying theoretical assumptions, will yield a reasonable approximation to causal estimates of the desired estimands (i.e., the treatment effects). Moreover, even if implementing a strict marginal structural model is not feasible, the IPTW framework remains a valid and desirable approach, as it overcomes many limitations often encountered by other causal frameworks [23].

The weights for binary exposures will be estimated through binomial logistic regression. Similarly, the weights for multicategory, nominal treatments (e.g., LRT modality) will be estimated using multinomial logistic regression. Where possible, in all analyses, baseline covariates will include patient BMI, age, gender, ethnicity, diagnosis of diabetes, functional status (ECOG performance score), Child Pugh score classification, MELD score, graft status, and donor age, ethnicity, gender, and BMI. Additionally, for LRT analyses, baseline covariates will include the Milan Criteria for Liver Transplantation and Alpha-Fetoprotein. In estimating the IPTWs, various model specifications will be explored, guided by clinical knowledge (i.e., to inform interactions and higher-order terms). The final model (among the set of candidate models) will be chosen with respect to the Akaike Information Criterion [26] while endeavouring to minimize overfitting and using clinical judgement for realistic interactions or higher-order terms.

Treatment effects for post-transplant survival, continuous outcomes, nominal categorical outcomes, and ordinal categorical outcomes will be estimated by multivariable cox regression, ordinary least squares regression, binomial or multinomial logistic regression, and cumulative logistic regression (i.e., proportional odds), respectively, each in the MSM framework.

In addition to the model specification for the exposure model, model specifications for the outcome model will be investigated. Additionally, the impact of truncating the derived weights at varying percentiles will be explored, ultimately endeavouring to balance bias and variance. All analyses will be conducted in R (version 4.01 or higher). For each effect estimate, as per convention, we will report upon the point estimate, the asymptotic 95% confidence interval, and corresponding p-values.

## DISCUSSION

### Clinical practice and implications

Currently, there is no level 1 evidence regarding the clinical indications for use and long-term effects of TIPS in the pre-orthotopic liver transplant setting. While some prior studies have pointed towards a beneficial survival effect with pre-transplant TIPS, the effect has been demonstrated to be modest [27, 28] and there are conflicting reports of this effect. Similarly, there are studies that support pre-transplant bridging-LRT with improved recurrence and survival [29, 30]; however, there are numerous similar studies with converse findings or findings of no effect [31-34].

It is critically important to clearly understand the pre-OLT effect of TIPS on overall and post-OLT specific survival intervals to assess its overall therapeutic role in the treatment of pre-transplant end-stage liver disease patients. This study may provide evidence to further support its use in the existing clinical setting or change its priority in the treatment paradigm as even a prophylactic rather than acute and reactive intervention. These findings may differ among those with and without hepatocellular carcinoma, which is accounted for in the study methods.

Patients with HCC may receive curative or bridging locoregional therapy (LRT) based on tumor characteristics and staging, which involve use of the Barcelona-Clinic Liver-Cancer (BCLC) staging system [35]. The BCLC staging system provides guidance on intervention types, which include chemotherapeutic, surgical, and locoregional interventions. Locoregional therapies can broadly be categorized as intra-arterial, percutaneous, and non-invasive (i.e., external beam radiation). While not exhaustive in the clinical definition of LRTs, the above categorization may provide valuable insights into the therapeutic benefits (in clinical and survival outcomes), both relative and absolute, to candidate patients. The results of this large database retrospective study may better inform existing clinical recommendations or lay the groundwork for further clinical investigation (i.e., prospective studies).

### Strengths and Limitations

This will be, to our knowledge, the first population-based study to analyze the effect of TIPS in OLT patients. Moreover, it will be the first study to causally analyze the effect of TIPS and the effect of LRTs in OLT patients with HCC. Linking the UNOS datasets will provide a unique chance to rigorously evaluate patient clinical status and survival outcomes in the perioperative and post-OLT setting. This will provide much-needed insight into the clinical impact and survival of TIPS with and without LRT in both HCC and non-HCC populations. This study will allow interventions to be compared at various stages in the patient’s clinical pathway, namely, perioperatively and postoperatively. Cumulatively, the results of this study may be the starting point for evidence-based care among this at-risk population and provide a basis for the design of future prospective studies.

Despite the clear importance of the proposed study, it is not without limitations.

- It is a retrospective study, suggesting that covariates may be inadequately reported, and if reported, may not align with necessary time scales; we have touched upon this above.
- Like any observational study, there exists the possibility of unmeasured confounding, which poses limitations on the reliability of estimated causal effects. While this is neither entirely verifiable (by definition), our study protocol aims to capture the most relevant clinical information, and where possible, we will make use of additional information from the UNOS database.
- In the analyses of the locoregional therapies in HCC patients, the sample size for a given intervention may be relatively small, thus yielding underpowered and inefficient estimates in the corresponding analyses. This issue may be exacerbated and persist across other analyses if a complete-case analysis is conducted for missing data (depending on the considerations discussed above).

## ETHICS AND DISSEMINATION

This study protocol was reviewed by the Emory University School of Medicine Institutional Review Board (IRB), and ethical approval was waived due to the retrospective analysis of the originally anonymized database. Disseminations of the proposed research will include peer-reviewed publications, conference presentations, and social media. Additionally, the results of the proposed research may be disseminated at a continuing medical education seminar. It was not appropriate or possible to involve patients or the public in the design, or conduct, or reporting, or dissemination plans of our research.

## Data Availability

Data is provided by UNOS database.

## Acknowledgements

The data reported here have been supplied by the United Network for Organ Sharing as the contractor for the Organ Procurement and Transplantation Network.

## Author Contributions

MK: Designed the study, drafted the manuscript, critically reviewed the manuscript

JTM: Designed the study, drafted the manuscript, critically reviewed the manuscript

PH: Critically reviewed the manuscript

HSK: Helped with planning of the study and critically reviewed the manuscript

MX: Helped with planning of the study

NN: Designed and supervised the study, drafted the manuscript, and critically reviewed the manuscript

